# FA-NIVA: A Nextflow framework for automated analysis of Nanopore based long-read sequencing data for genetic analysis in Fanconi anemia

**DOI:** 10.64898/2026.02.27.26346867

**Authors:** Priya Satish Neurgaonkar, Michelle Dierolf, Luke O’Gorman, Christian Remmele, Judith Schäffer, Isabell Popp, Angela Borst, Simone Rost, Markus J. Ankenbrand, Christian P. Kratz, Anke K. Bergmann, Reinhard Kalb, Jiangyan Yu

## Abstract

**Motivation:** Fanconi anemia (FA) is a rare disease mainly caused by biallelic pathogenic variants, including structural variants such as large deletions and insertions in FA genes. Currently, variant detection is based on short-read sequencing and probe-based approaches. However, determining the exact genomic breakpoint or achieving allelic discrimination remains challenging. Nanopore-based long-read sequencing enables a comprehensive detection of FA variants, but a unified bioinformatic analysis platform for these data is missing.

**Results:** We present FA-NIVA (<u>F</u>anconi <u>a</u>nemia - <u>N</u>anopore <u>I</u>ndel and <u>V</u>ariant <u>A</u>nalysis), an automated and adaptable analysis workflow tailored for Nanopore-based long-read sequencing data in FA genetic analysis. FA-NIVA integrates state-of-the-art tools to comprehensively detect both single nucleotide variants (SNVs) and structural variants (SVs). Our analysis platform enhances genotyping accuracy for biallelic variants by a joint SNV-SV based phasing in FA associated genes. Built within the Nextflow ecosystem and powered by containerized Docker images, FA-NIVA ensures reproducibility, flexibility, scalability and transparency across different computing environments. Together, FA-NIVA provides a robust end-to-end solution for the automated analysis of SVs and SNVs and high-resolution phasing analysis in FA genes, enabling an accurate and efficient pipeline for genetic analysis.

**Availability:** FA-NIVA is available on GitHub at: https://github.com/UKWgenommedizin/FA-NIVA.

## 1 Introduction

Fanconi anemia (FA) is a rare inherited disorder caused by pathogenic variants in one of at least 22 FA-associated genes (Kuehl et al., 2025). Most variants follow an autosomal recessive inheritance, except for *FANCB* (X-linked) and *FANCR* (autosomal-dominant). About 60% of FA patients harbor biallelic variants in *FANCA* (Stenson et al., 2020; Mehta et al., 2021), with up to 23% involving large deletions spanning over 545 kb (Levran et al., 2005; Ameziane et al., 2008; Moghrabi et al., 2009; Kimble et al., 2018). These variants often occur in *Alu-*rich regions. Another gene, *FANCD2*, is flanked by pseudogenic regions, which complicates the detection of gene specific variants (Timmers et al., 2001; Kalb et al. 2007; Shangguan et al., 2025).

Although short-read sequencing is generally highly reliable to detect single nucleotide variants (SNVs) and indels within the coding sequence, it is challenging to detect complex or large structural variants (SV) or to resolve repetitive sequences. Multiplex ligation-dependent probe amplification (MLPA) of *FANCA* provides accurate information on copy numbers. However, determination of the precise genomic breakpoint requires multiple complementary molecular approaches. Another challenge is the allelic assignment of variants, which is required for confirming compound-heterozygosity. Consequently, SVs are frequently incompletely characterized or require the combination of various methods; confirmation of compound-heterozygosity relies largely on allelic segregation analyses.

Long-read sequencing (LRS) technologies generate read lengths ranging from several kb to over 10 kb, enabling simultaneous detection of clinically relevant SNVs and SVs for comprehensive variant profiling in diagnostics (Sen et al., 2025). Despite these advances, broader clinical implementation of LRS in routine diagnostics remains limited by the lack of standardized and automated analysis workflows. Existing pipelines, such as the Nextflow workflow nanoseq (https://github.com/nf-core/nanoseq), offer general-purpose processing components for Nanopore sequencing data, but are not designed for complex variant detection. Moreover, these pipelines do not incorporate phasing analysis for autosomal recessive disorders, lacking the confirmation of biallelic occurrence of pathogenic variants. Implementation of LRS in routine diagnostics for Fanconi anemia requires the development of a reliable, transparent and easy-to-use toolset with options for customizability.

We present FA-NIVA (<u>F</u>anconi <u>a</u>nemia – <u>N</u>anopore <u>I</u>ndel and <u>V</u>ariant <u>A</u>nalysis), a robust and modular-based bioinformatic pipeline for the joint detection of SNVs and SVs, as well as phasing and genotype correction within deletions. The pipeline is specifically designed for FA-genes that contain challenging regions, such as high frequency of *Alu*-elements and pseudogenes. FA-NIVA integrates automated QC and provenance reporting to ensure diagnostic-grade traceability, while its modular architecture provides flexibility required for diverse clinical applications.

## 2 Feature overview

FA-NIVA performs end-to-end data analysis from raw signal processing and read alignment to variant calling, with additional modules for phasing and variant annotation (Figure 1). Below highlights the key features of the pipeline.

**Figure 1.**
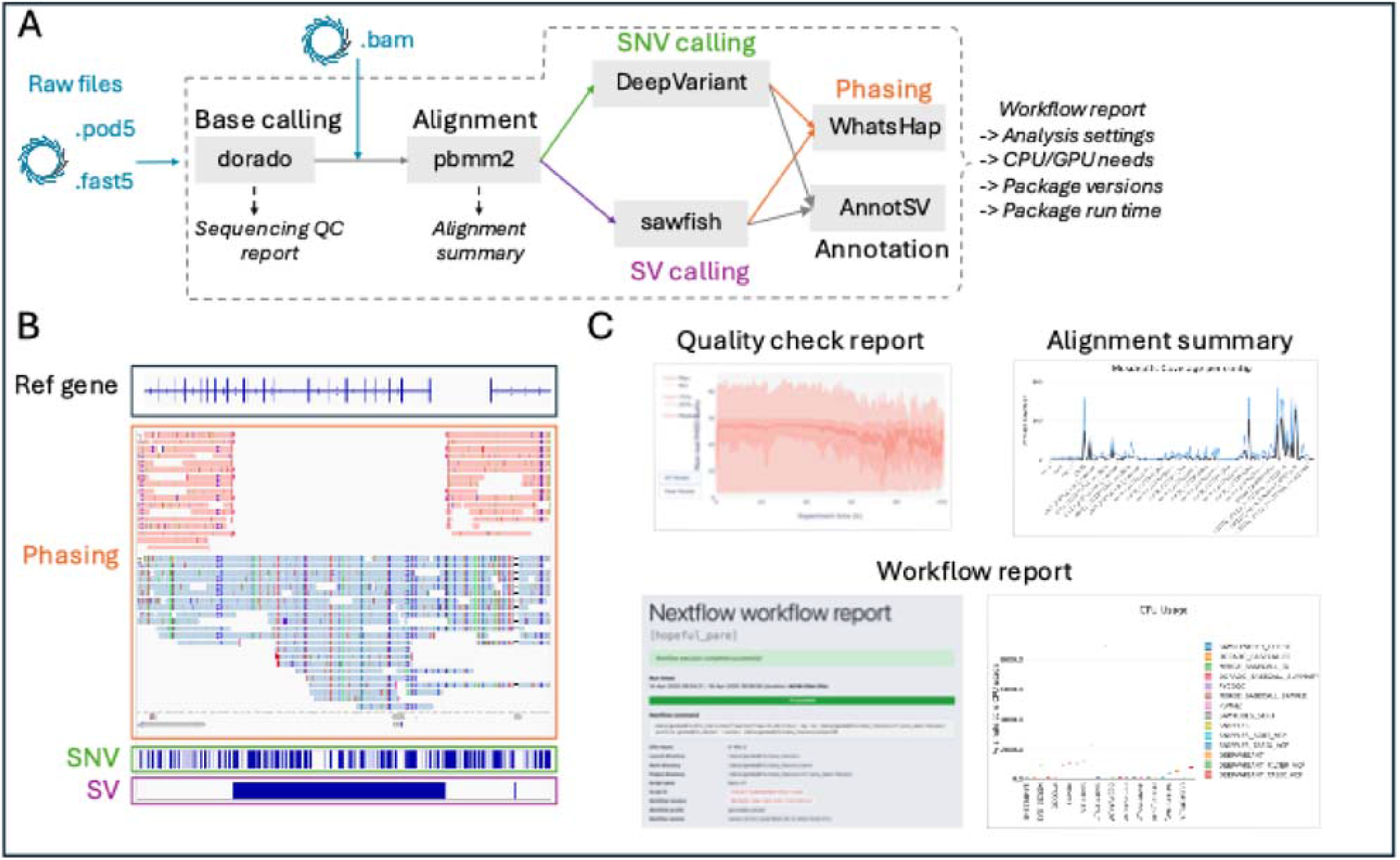
Overview of the FA-NIVA pipeline and key features. A Schematic overview of tools integrated in FA-NIVA. Analysis tools integrated in FA-NIVA are highlighted in grey boxes, with corresponding analysis steps annotated above/below each tool. Reports generated at each step are shown in italics. B Visualization of identified variants in Integrative Genomics Viewer (IGV). C Workflow reports automatically generated by the pipeline.

### Support for multiple input file types

FA-NIVA accepts all standard raw file formats generated by Nanopore sequencers, including .pod5, .fast5 and previously generated .bam. Upon upload, electrical signals are automatically converted into the corresponding nucleotide sequences using the Dorado basecaller on a computing system equipped with GPUs. The resulting bam file is then seamlessly transferred to variant calling modules for SNVs and SVs (Figure 1A).

### High-confidence variants calling analysis

FA samples frequently harbour large *FANCA* deletions with near-identical flanks, challenging a correct breakpoint placement for downstream variant calling analysis. Minimap2 is the recommended aligner for Nanopore sequencing data and offers a high overall accuracy (Helal et al., 2024). However, we observed critical errors in handling of soft-clipped sequences in supplementary alignment for reads containing large deletions (Supplementary Figure 1), leading to incorrect genomic position in subsequent SV calling. In contrast, pbmm2, the minimap2 wrapper, is able to accurately align reads across large deletions. This is likely because pbmm2, tuned for high quality HiFi reads with refined heuristics, anchors reads more reliably. Standard minimap2, optimized for noisier Nanopore sequencing data, might overcompensate for sequencing errors and misplace breakpoints. Therefore, based on our experience with FA data analysis, we integrated pbmm2 (preset -CCS) into our pipeline, which provides most accurate alignment results and improves SVs detection.

FA-NIVA integrates high-performance tools for variant calling. Our in-house benchmarking of SNV calling on Nanopore sequencing data demonstrates that DeepVariant (Poplin et al., 2018) outperforms other tools, achieving an F1 score of 0.998. The most recent SV caller, sawfish, extracts read sequences near breakpoint regions to generate consensus contigs, improving the accuracy of breakpoint determination for SVs (Saunders et al., 2025). Using this refined cluster-based approach, we also found that sawfish can precisely identify deletion breakpoints in regions with nearly identical flanking sequences, where accurate alignment remains a significant challenge for existing algorithms. Therefore, by combining DeepVariant for SNV detection and sawfish for SV calling, FA-NIVA ensures high-confidence detection of clinically relevant variants, which is crucial for a comprehensive toolkit.

Automatic variant annotation and ranking are performed using AnnotSV (Geoffroy et al., 2023). The annotation provides both fundamental information including gene names, regulatory elements and breakpoint details, as well as clinically relevant data. The annotated variant list provides a robust framework for downstream filtering and variant interpretations.

### High-resolution phasing analysis via joint SNV-SV

Accurate phasing is crucial for the analysis of compound heterozygosity and for distinguishing between homozygous and hemizygous variants in regions where one allele harbours a large deletion. However, most current phasing algorithms use only SNVs, which can lead to inaccurate inference of allele inheritance (Martin et al., 2023). Specifically, we identified incorrect haplotagged reads arising from SNVs within large deletions that were misclassified as homozygous rather than hemizygous (Supplementary Figure 2A). Simply combining SNVs and SVs has not been sufficient to improve haplotagging accuracy (Mahmoud et al., 2021). FA-NIVA introduces a genotype-aware correction strategy that explicitly incorporates SVs into the phasing process. By systematically correcting the genotypes of SNVs located within large deletions, mis-haplotagging can be avoided and reliable zygosity information can be provided (Figure 1B and Supplementary Figure 2B). Furthermore, large SVs, which were previously excluded from phasing due to their sparse distribution and lower genotype confidence, can be used to improve the resolution in our pipeline.

### Detailed performance summary for analysis run

FA-NIVA generates a comprehensive workflow report, a sequencing summary from copyQC (Leger et al., 2019), and alignment summaries produced by MultiQC (Ewels et al., 2016) and mosdepth (Pedersen et al., 2018) (Figure 1C). The report also contains: i) the exact command-line input and analysis parameters used, ii) resource usage such as CPU/GPU allocation and run time for each step, iii) the Docker image version used for each module, and iv) the execution time for individual steps. This detailed information ensures transparency and reproducibility enabling users to replicate or troubleshoot their analyses.

## 3 Dissemination and use-cases

To ensure broad accessibility and transparency, the source code of FA-NIVA package is deposited at GitHub: https://github.com/UKWgenommedizin/FA-NIVA. Extensive instructions for installing, configuring, and running the FA-NIVA are provided in the GitHub repository, along with documentation of input requirements, parameter options, and example/test datasets to guide users through practical implementation.

To demonstrate the performance and flexibility of FA-NIVA, three use-cases using different input file types with selected analysis modules are shown in the supplementary file:

### Use-case 1: Detection of bi-allelic *FANCA* variants including a large deletion (POD5 input)

The workflow accurately determined previously reported bi-allelic variants in an FA sample, including the precise determination of a 77.8kb deletion (QUAL of 251, QUAL = −10 × log10 (Probability (call is wrong))) spanning *FANCA* gene exon 3 and extending beyond the 3’-UTR region, and one SNV within the *FANCA* noncoding region (Supplementary Figure 3). This resolves the MLPA-only detection of this deletion by providing nucleotide-level breakpoints.

### Use-case 2: Identification of an insertion of an *Alu* element in the *FANCD2* gene (POD5 input)

Using raw signal-level POD5 files, the workflow detected a high quality (QUAL of 504) 299bp insertion event within the *FANCD2* gene (Supplementary Figure 4). This confirms that insertions, also from highly repetitive elements of approximately 300 bp in size, are reliably recognized with the integrated SV caller.

### Use-case 3: Demonstration of broader applicability: Identification of large homozygous region in a patient with limb-girdle muscular dystrophy, autosomal recessive 2 (BAM input)

Chromosome 2 appears heterozygous, while FA-NIVA identified a homozygous region spanning 227 kb (chr2:71,241,093-71,468,910), located in the 5’-UTR region of the *DYSF* gene (Supplementary Figure 5). True homozygosity was confirmed by phasing of the reads. Secondary validation was reached by short read whole genome sequencing using a NovaSeq 6000 system (Illumina). This illustrates that the workflow can be repurposed for non-FA recessive disorders that benefit from phasing.

## 4 Conclusion

FA-NIVA addresses several key bioinformatic bottlenecks that currently limit routine analysis of datasets generated by Nanopore sequencing. By automating the entire process from raw sequencing signal handling to variant calling, FA-NIVA substantially reduces manual intervention. It robustly supports all file formats generated by the Nanopore platform and provides a comprehensive toolkit that reliably detects both small SNVs and large SVs, while simultaneously delivering accurate phasing information through its integrated SNV-SV toolset. Built within the Nextflow ecosystem (Di Tommaso et al., 2017), the containerized analysis modules ensure reproducibility across different computing environments. Furthermore, the pipeline generates detailed reports that document the analysis environment and provides a clear, first-glance overview of data quality and results. This end-to-end automation ensures consistency, transparency and efficiency at diagnostic standards.

Our use-cases demonstrate that FA-NIVA is not only able to identify canonical variants but also to resolve structural variants. Moreover, FA-NIVA enables accurate allelic discrimination for precise SNV annotation, confirmation of compound-heterozygosity, and detection of homozygous regions. Due to its modular architecture, the pipeline can be easily adapted to specific tasks, such as homozygosity mapping in consanguineous families or analysis of alternative data types (e.g. PacBio data). One module not yet implemented is methylation analysis, which will be incorporated in future versions.

Overall, FA-NIVA provides an automated bioinformatics pipeline for indel and variant analysis of Nanopore-based sequencing data in FA-genes. Accurate phasing allows reliable data interpretation. Moreover, the customizable and module-based architecture enables robustness and adaptability for broader applications across genetic disease datasets.

## Data Availability

All data produced are available online at GitHub

https://github.com/UKWgenommedizin/FA-NIVA

## Acknowledgements

We would like to thank Joshua Bopp, Madita Huvar and Dr. Jonny Derk for the technical support in setting up the sequencing runs and Dr. Claudia Davenport for revising the manuscript.

## Contribution

Conceptualization of the study: A. Bergmann, J. Yu & R. Kalb. Pipeline development: P. S. Neurgaonkar, L. O’Gorman, C. Remmele, M. J. Ankenbrand & J. Yu. Sequencing and validation: P. S. Neurgaonkar, M.

Dierolf, J. Schäffer, I. Popp, A. Borst, S. Rost & R. Kalb. Supervision: R. Kalb. Writing: P. S. Neurgaonkar, M. Dierolf, R. Kalb & J. Yu.

