## Supplementary Information for "FA-NIVA: A Nextflow framework for automated analysis of Nanopore based long-read sequencing data for genetic analysis in Fanconi anemia"

Supplementary methods

The FA-NIVA pipeline can be downloaded from github.

git clone -b main https://github.com/UKWgenommedizin/FA-NIVA

First, edit the samplesheet.csv file to specify the input data paths.

id,sample,flowcell,input_path,batch,kit

run31,run31_supmode,flowcell,/home/raw_data/run31/pod5,20250402,LSK114

run30,run30_supmode,flowcell,/home/raw_data/run30/pod5,20250402,LSK114

run29,run29_supmode,flowcell,/home/raw_data/run29/pod5,20250402,LSK114

Then, modify the profile.config file to define the reference data directory. Update the file paths accordingly to match your local system.

```
params {
    config_profile_name        = 'Fanconi profile'
    config_profile_description = 'Analyze fanconi nanopore dataset'
    // Input data for minimal size test
    input                      = "${baseDir}/assets/samplesheet.csv"
    // Genome reference
    fasta                      = "/data/genmedbfx/ref/GRCh38.fasta"
    fasta_index                = "/data/genmedbfx/ref/GRCh38.fasta.fai"
    // Regions to modify SNV genotypes for phasing step
    SNV_modify_regions         = "${baseDir}/assets/SNV_modify_regions.csv"
    //annotsv annotation direcoty
    annotsvAnnotationsDir      = "/data/genmedbfx/ref/annosv/"
    dorado_model               = 'dna_r10.4.1_e8.2_400bps_fast@v5.0.0'
    dorado_files_chunksize     = 10000
    dorado_modifications_model = ''
    reads_format               = 'bam'
}
```

Next, adjust the nextflow.config file to indicate which modules to include in the analysis, by setting each option to true or false.

The last step is to configure computational resources in the base.config file. By default, the pipeline uses one GPU core for dorado basecaller and deepvariant steps. Assigning additional GPUs may lead to the error of “CUDA out-of-memory”.

Finally, run the pipeline using the command shown below. The resulting output files will be written to the directory specified in the configuration. An example workflow report is available on Zenodo: <https://zenodo.org/records/17284961>

Nextflow run ./FA-NIVA/ \ # The path to the FA-NIVA package

-profile fa_niva,docker \

--outdir ./output

Supplementary Figures

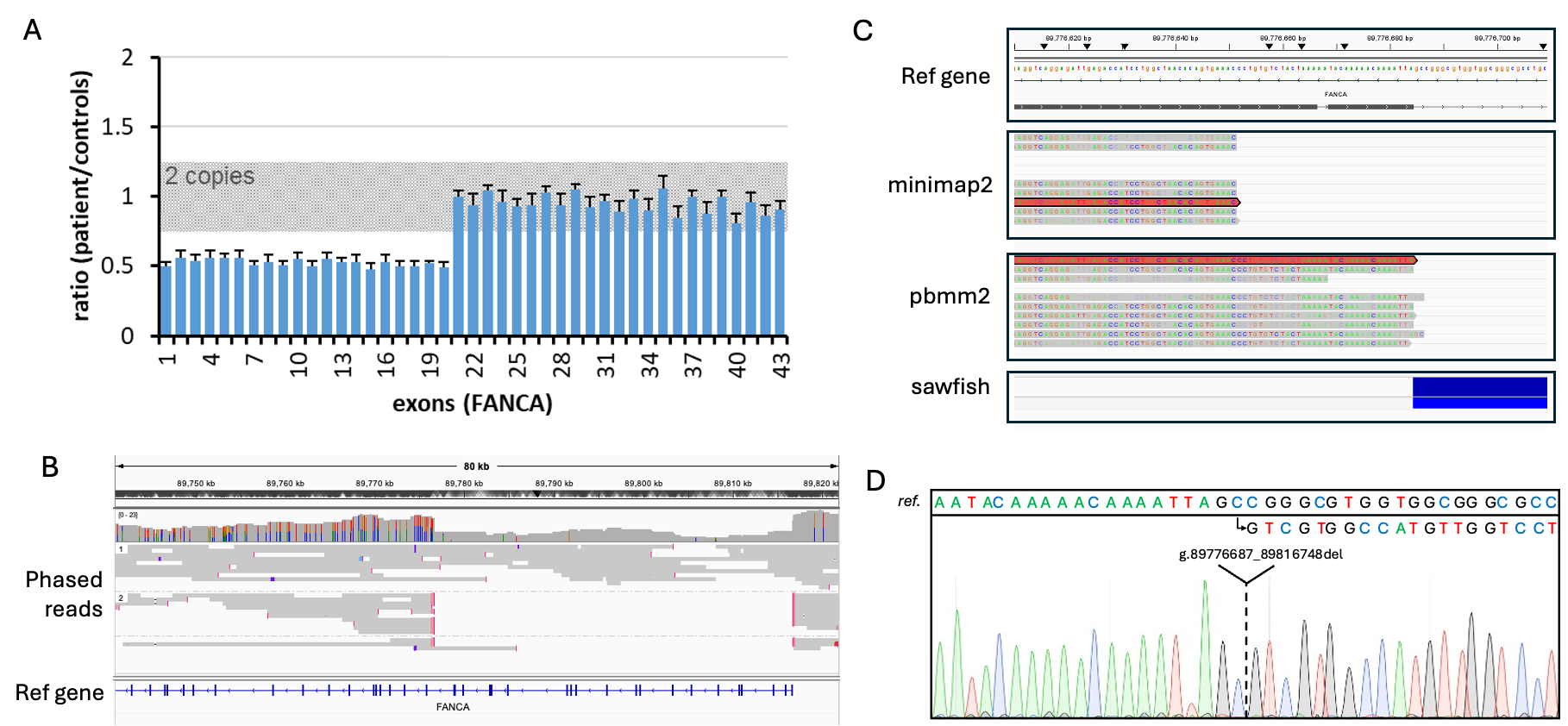

Supplementary Figure 1. The pbmm2 based alignment solves soft-clipped sequences at the deletion breakpoint. **A** A large heterozygous deletion spanning *FANCA* exon 1 to exon 20 was identified in this sample by Multiplex Ligation-dependent Probe Amplification (MLPA). **B** FA-NIVA pipeline successfully determined this 40kb deletion at chr16:89776684-89816746 (QUAL of 395, QUAL = -10 × log10 (Probability (call is wrong))) encompassing exons 1 through exon 20 of the *FANCA* gene. The variant is described according to Human Genome Variation Society (HGVS) nomenclature as NC_000016.10:g.89776687_89816748del. **C** Comparison of minimap2 and pbmm2 alignments at the breakpoint of the large deletion. The large deletion detected by sawfish is shown at the bottom. The minimap2 based alignment incorrectly clipped a 33 bp sequence, while the pbmm2 based alignment correctly soft-clipped this region, improving breakpoint resolution. **D** Sanger validation of the breakpoint.

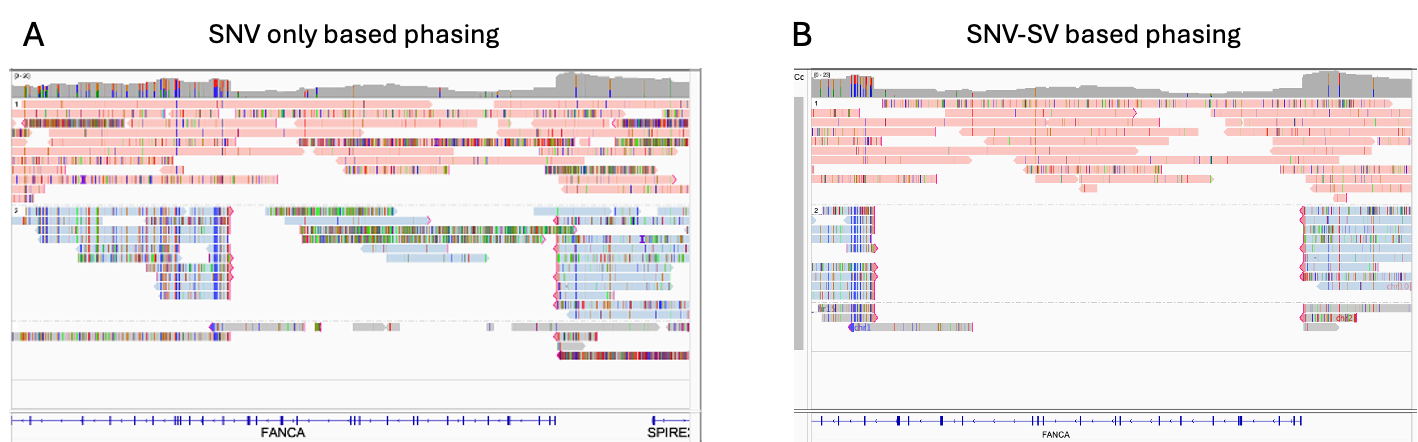

Supplementary Figure 2. Integrating both SNVs and SVs in phasing analysis enables accurate haplotagging of reads. **A** SNVs only based analysis failed to haplotag all reads. **B** Joint SNV-SV phasing analysis accurately haplotag all reads.

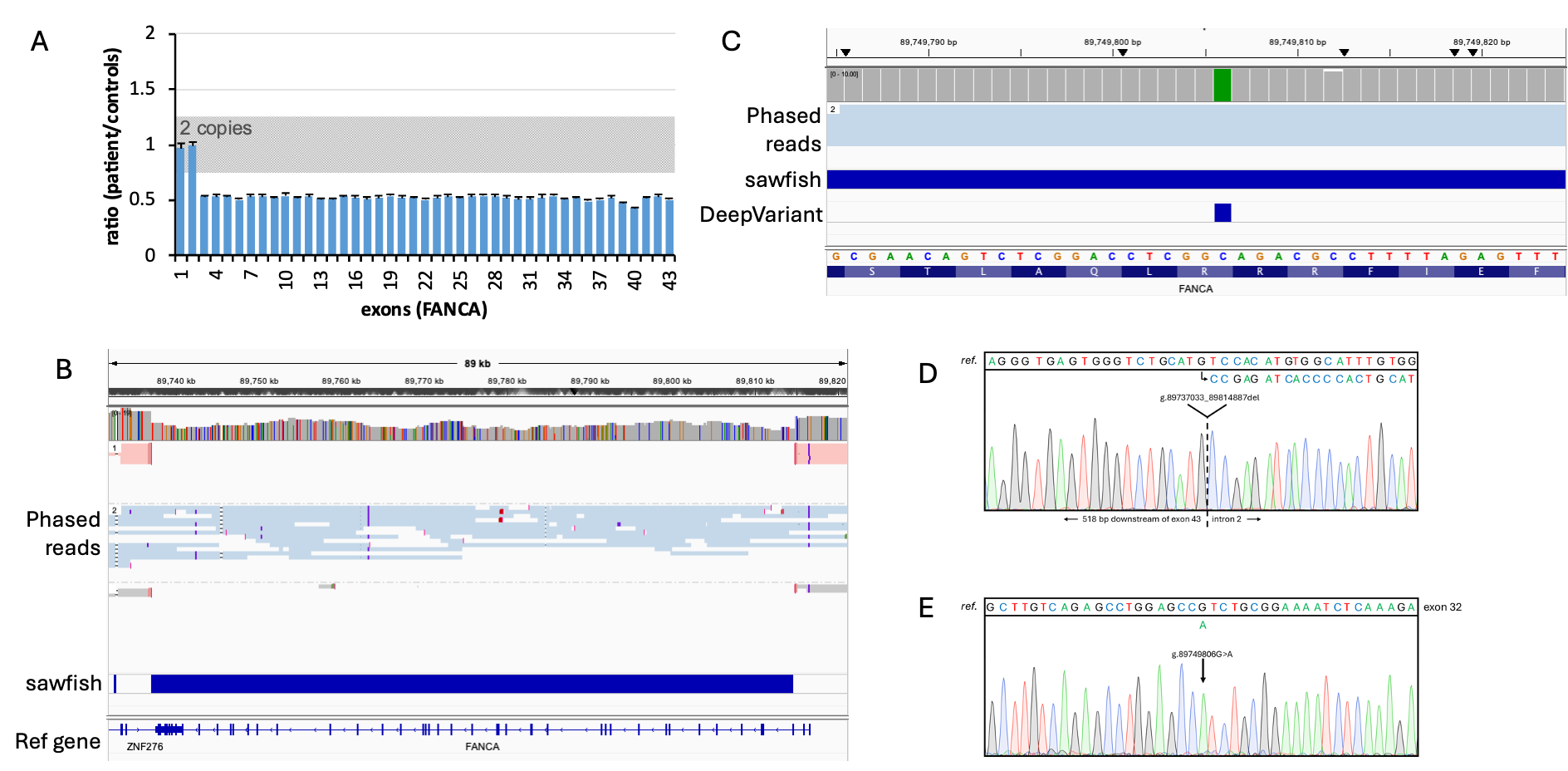

Supplementary Figure 3. Two bi-allelic variants including one 77.8kb deletion and one SNV were detected in one FA sample. **A** A large deletion spanning *FANCA* exon 3 and exceeding 3’-UTR was identified in this sample by MLPA. **B** The FA-NIVA pipeline determined this 77.8kb deletion at chr16:89737031-89814886 (QUAL of 251) on one allele (colored reads in pink). The variant is described according to HGVS nomenclature as NC_000016.10:g.89737033-89814887del. **C** The FA-NIVA pipeline determined the SNV NC_000016.10:g.89749806G>A on another allele (reads colored in blue). **D** Sanger validation of the breakpoint. **E** Sanger validation of the SNV.

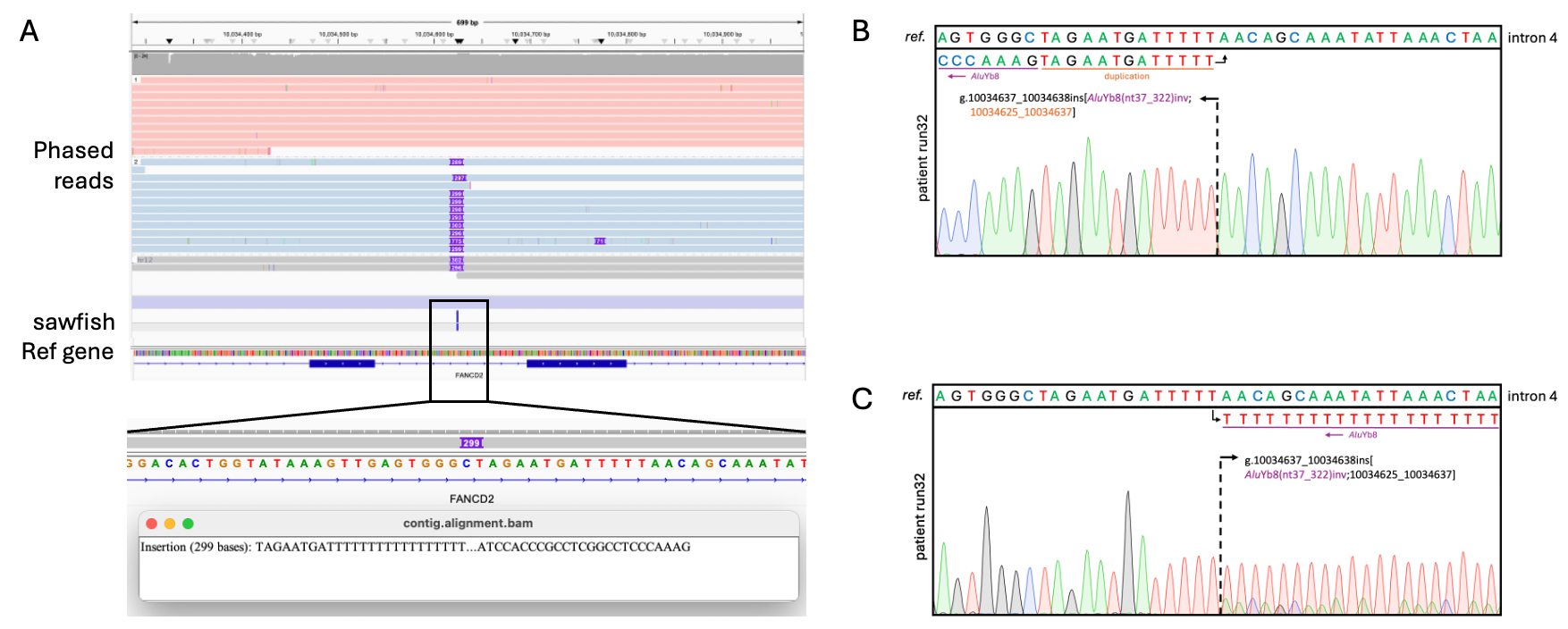

Supplementary Figure 4. Insertion of an *Alu* element in the *FANCD2* gene was determined in one FA sample. **A** FA-NIVA pipeline determined this 299bp insertion at chr3:10034624 in one allele with a QUAL score of 504. The variant is described according to HGVS nomenclature as NC_00003.12:g.10034637_10034638ins[*Alu*Yb8(nt37_322)inv;10034625_10034637]. **B** Sanger validation of the insertion by forward primer. **C** Sanger validation of the insertion by reverse primer.

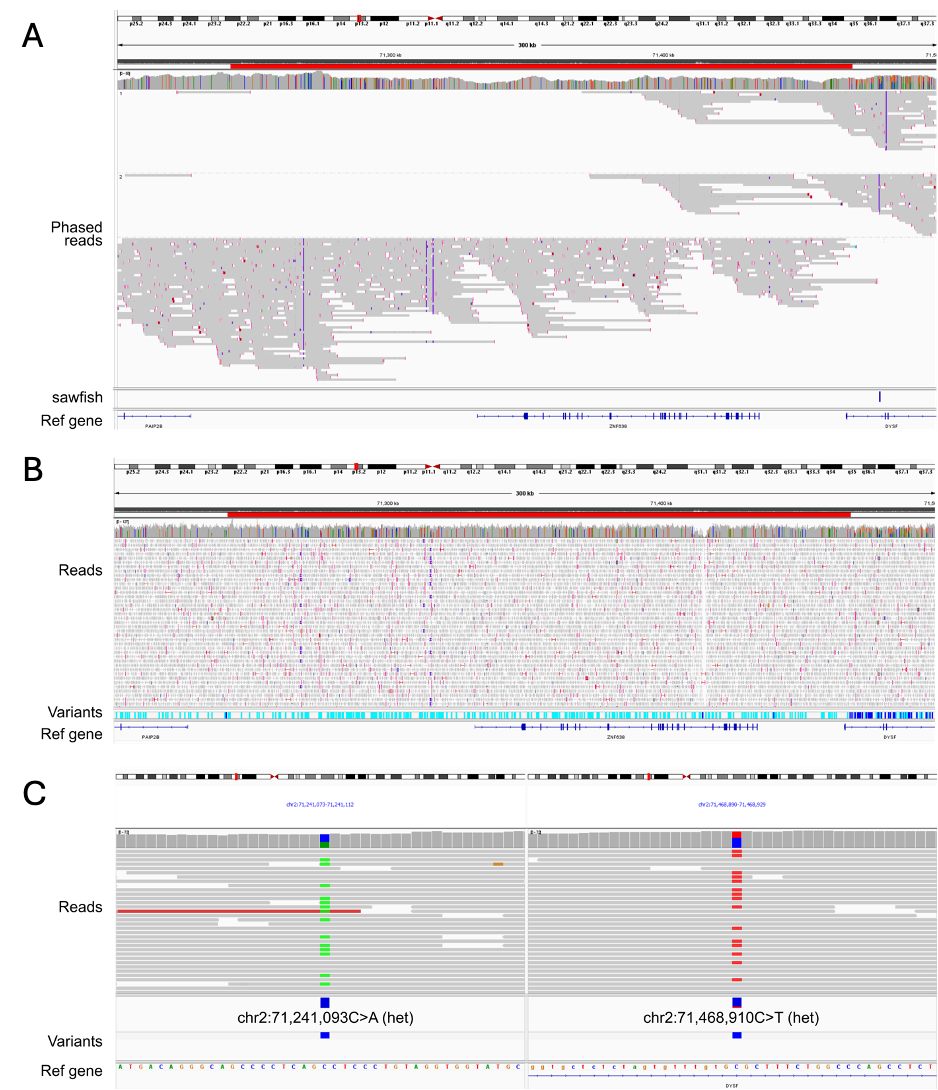

Supplementary Figure 5. A large homozygous region was determined in one LGMRD2 sample. **A** FA-NIVA pipeline determined a 227 kb homozygous region in the 5’UTR region of *DYSF* (chr2:71,241,093-71,468,910; region indicated with a red bar). **B** Short-read whole-genome sequencing validation of homozygous region (region indicated with a red bar). Homozygous variants are indicated in light blue; heterozygous variants are indicated in dark blue. **C** Short-read whole-genome sequencing validation of the borders of the homozygous region, due to heterozygous variants. Both were also observed in the long-read whole-genome sequencing data.
